# The Relationship of Socioeconomic Status, Antenatal Care Visits, and Hospital Delivery in Bangladesh: Analysis of Demographic and Health Survey 2022

**DOI:** 10.1101/2025.03.18.25324232

**Authors:** Gulam Muhammed Al Kibria, Md Shajedur Rahman Shawon, Mohammad Nurunnabi, Md Zabir Hasan

## Abstract

Bangladesh and many other low- and middle-income countries experience a high number of maternal and neonatal deaths; antenatal care (ANC) and hospital delivery are crucial to reducing these deaths. In this cross-sectional study, we investigated the relationship of socioeconomic status with at least 4 ANC visits and hospital delivery in Bangladesh. We also tested whether antenatal care mediated the association of socioeconomic variables and facility delivery. We used data from the Bangladesh Demographic and Health Survey 2022. After descriptive analysis, generalized structural equation modeling was used to investigate the associations.

A total of 4,950 women were included in the analysis (mean age: 25.7 years, 73.2% rural). The proportion of women with at least 4 ANC visits and hospital delivery was 39.8% and 64.4%, respectively. In adjusted analyses, all socioeconomic variables had significant associations with at least 4 ANC visits and hospital delivery. For instance, compared to women with the poorest wealth quintile, those with poorer (adjusted odds ratio (AOR): 1.26, 95% confidence interval (CI):1.02-1.55), middle (AOR: 1.43, 95% CI: 1.16-1.77), richer (AOR: 1.98, 95% CI: 1.59-2.45), and richest (AOR: 3.12, 95% CI: 2.45-3.99) wealth quintiles higher odds of at least 4 ANC visits. Similarly, for hospital delivery, compared to women with the poorest wealth quintile, those with poorer (AOR: 1.44, 95% CI: 1.19-1.75), middle (AOR: 1.82, 95% CI: 1.48-2.24), richer (AOR: 2.48, 95% CI: 1.98-3.10), and richest (AOR: 3.90, 95% CI: 2.94-5.18) wealth quintiles higher odds. Women with at least 4 ANC visits, had more than two times higher odds of hospital delivery (AOR: 2.56, 95% CI: 2.20-2.97). When we looked into the mediation, at least 4 ANC visits mediated 60.5%, 34.6%, and 41.1% of the relationships of women’s education, husband’s education, and household wealth with hospital delivery, respectively.

Considering the lower utilization of at least 4 ANC visits and its mediating impact on the relationship between socioeconomic status and facility delivery, more community-based programs are required to increase awareness about at least 4 ANC visits and hospital births.

## Introduction

Globally, with a disproportionately high maternal mortality ratio (MMR) and neonatal mortality rate (NMR), most maternal and neonatal deaths occur in low- and middle-income countries (LMICs) [1,2]. An estimated 287,000 women worldwide lost their lives to preventable pregnancy and childbirth-related causes in 2020, resulting in an MMR of 134 per 100,000 live births (LB) [1]. This number translates to over 800 deaths every day of that year. Furthermore, an estimated 2.3 million children died globally in 2022, resulting in an NMR of 17 per 1,000 LB and 6,300 deaths each day. With an MMR of 123 per 100,000 live births in 2022 and an NMR of 17 per 1,000 LB in 2022, Bangladesh, an LMIC in South Asia, experiences high maternal and neonatal mortality. Two key objectives of the Millennium Development Goals (MDG) were lowering child mortality and enhancing maternal health [3]. In 2014, this nation’s under-five mortality rate dropped to 46 per 1,000 LB, meeting the MDG objective [2]. For maternal deaths, it ended with 176 per 100,000 LB in 2015, despite its goal of reaching the MMR target of 143 deaths per 100,000 LB within that year [1]. The sustainable development goals (SDGs) aim to reduce the MMR and NMR to 70 per 100,000 and 12 per 1000 LB, respectively. It will be challenging for this country to reduce the MMR and NMR to the targeted level.

It should be mentioned that the main causes of maternal and newborn fatalities can be avoided or successfully addressed with currently accessible or available treatment options [4–6]. For instance, the main causes of maternal mortality include severe hemorrhage, infection, high blood pressure (including pre-eclampsia and eclampsia), and problems during delivery [7]. The leading causes of death among newborns are perinatal asphyxia, sepsis (i.e., infections), and prematurity/low birth weight [8]. Adequate antenatal care (ANC) has been suggested as the main strategy to increase the use of treatment for these causes and lower the number of deaths [6,9,10]. To ensure the adequate number of ANC and childbirth in health facilities, beginning ANC during the first trimester of pregnancy is recommended by the World Health Organization (WHO) [6]. Although they advise having at least eight ANC visits, the Bangladeshi government currently recommends four ANC visits. Initiating ANC during the first trimester or having at least 4 ANC visits is crucial to ensure facility delivery. Giving birth in healthcare facilities can significantly reduce maternal, perinatal, and neonatal fatalities [11,12]. A properly equipped medical facility with a clean environment for the delivery and training providers can substantially reduce obstetric complications, preventing maternal and newborn deaths and consequences [13,14].

Properly equipped health facilities not only provide ANC services but also ensure safe institutional deliveries, addressing critical gaps in maternal and neonatal healthcare. In Bangladesh, hospitals and health institutions at all levels offer obstetric care services. While village doctors, community health clinics, sub-district (i.e., upazila) health complexes/sub-centers, NGO-based health facilities, and private hospitals/clinics primarily provide health care services in rural areas, medical college hospitals, private hospitals/clinics, and district-level hospitals are the most prevalent in urban areas. In both areas, pharmacies are important providers of healthcare [15]. According to the latest Bangladesh Demographic and Health Survey (BDHS) of 2022, only 41% of women received at least 4 ANC visits and about 65% of women give birth in health facilities [16]. This has been a substantial increase over the past two decades. For instance, according to BDHS 1993-94, only 6% of women received at least 4 ANC visits and only 4% of women had childbirth in health facilities [17]. Furthermore, studies conducted in Bangladesh and other comparable LMICs revealed socioeconomic differences in obstetric care, with women from lower socioeconomic backgrounds less likely than those from higher socioeconomic backgrounds to obtain adequate ANC or facility delivery [18,19]. For instance, a previous study by Nayeem et al. found more than 3 times higher odds of facility delivery among women with the richest household wealth quintile than those with the poorest household wealth quintile [20]. A similar association has been observed for adequate ANC visits [21]. Another socioeconomic variable, education level (i.e., of women or their husbands), had a similar association, and women with higher education or higher educated husbands had an increased likelihood of adequate ANC than those with lower education or lower educated husbands [20,21]. The Government of Bangladesh has also taken steps to increase the availability, accessibility, and affordability of obstetric services among women with low socioeconomic status [16].

With significant investments in maternal and child health in Bangladesh and other LMICs, more women benefit from comprehensive care during pregnancy and childbirth. While the increase in ANC uptake may lead to higher rates of institutional deliveries-supporting the supply side of healthcare delivery-the demand-side determinants, such as socio-cultural factors, economic barriers, and individual educational attainments, remain poorly understood. These underlying factors must be explored further, as they cannot be fully identified or addressed by simply proving the statistical correlation between ANC utilization and facility-based births, requiring an updated analysis focusing on the abovementioned demand-side determinants. Understanding the effects of adequate ANC on the demand dynamics of institutional births is crucial for designing interventions that effectively enhance maternal and neonatal health outcomes.

In this study, we investigated the socioeconomic disparities in at least four ANC visits and facility delivery in Bangladesh. We also investigated if at least four ANC visits mediate the relationship between socioeconomic status and facility delivery. This study will be helpful for researchers and policymakers to better understand the impact of socioeconomic status on them, how ANC visits positively impact facility delivery, and how to design programs and policies in light of new findings.

## Methods

### Study Design and Setting

This cross-sectional study used data from BDHS 2022. BDHS is a nationally representative survey in Bangladesh. With a land area of 55,000 square miles and an estimated 170 million people, Bangladesh is a South Asian nation. Data on Bangladesh’s primary demographic, maternal, and child health indicators were collected. All divisions, the nation’s biggest administrative units, were included in the survey. As part of the global DHS program, this is the eighth DHS in the country. Data was collected between June and December of 2022 [16].

First, a list of enumeration areas (EA) was created as part of a sample frame. The frame was based on the 2011 housing and population census of the country. The sampling frame was created to obtain distinct estimates for each administrative division, as well as for rural and urban areas. Household selection involved two phases. A total of 250 EAs were selected in metropolitan areas, and 425 EAs were selected from rural areas during the first stage. A complete listing of all households was done. In the next stage, a sample of 45 households was selected from each EA. Lastly, 30 of the 45 households were randomly selected for individual long interviews of women. The survey had a 98% response rate. Interviews were conducted among women in 30,375 households; 19710 and 10,665 households from rural and urban regions, respectively. Only women of reproductive age who have been married and are between the ages of 15 and 49 were included in this study. The final sample contains 4950 women, 3621 from rural and 1329 from urban regions. The survey report, methodology, sample size calculation, and questionnaires are available online [16].

### Outcomes

This study has two primary outcomes: (a) the adequate number (i.e., at least four) of ANC visits, as recommended by the Government of Bangladesh; and (b) facility delivery, any delivery that happened at a hospital was regarded as a facility delivery. Home delivery was defined as any delivery that happened at their house, a friend’s house, or a relative’s house. The adequate number (i.e., at least four) of ANC visits was also investigated as a mediator in the relationship between socio-economic status and facility delivery (Figure 1).

**Figure 1:**
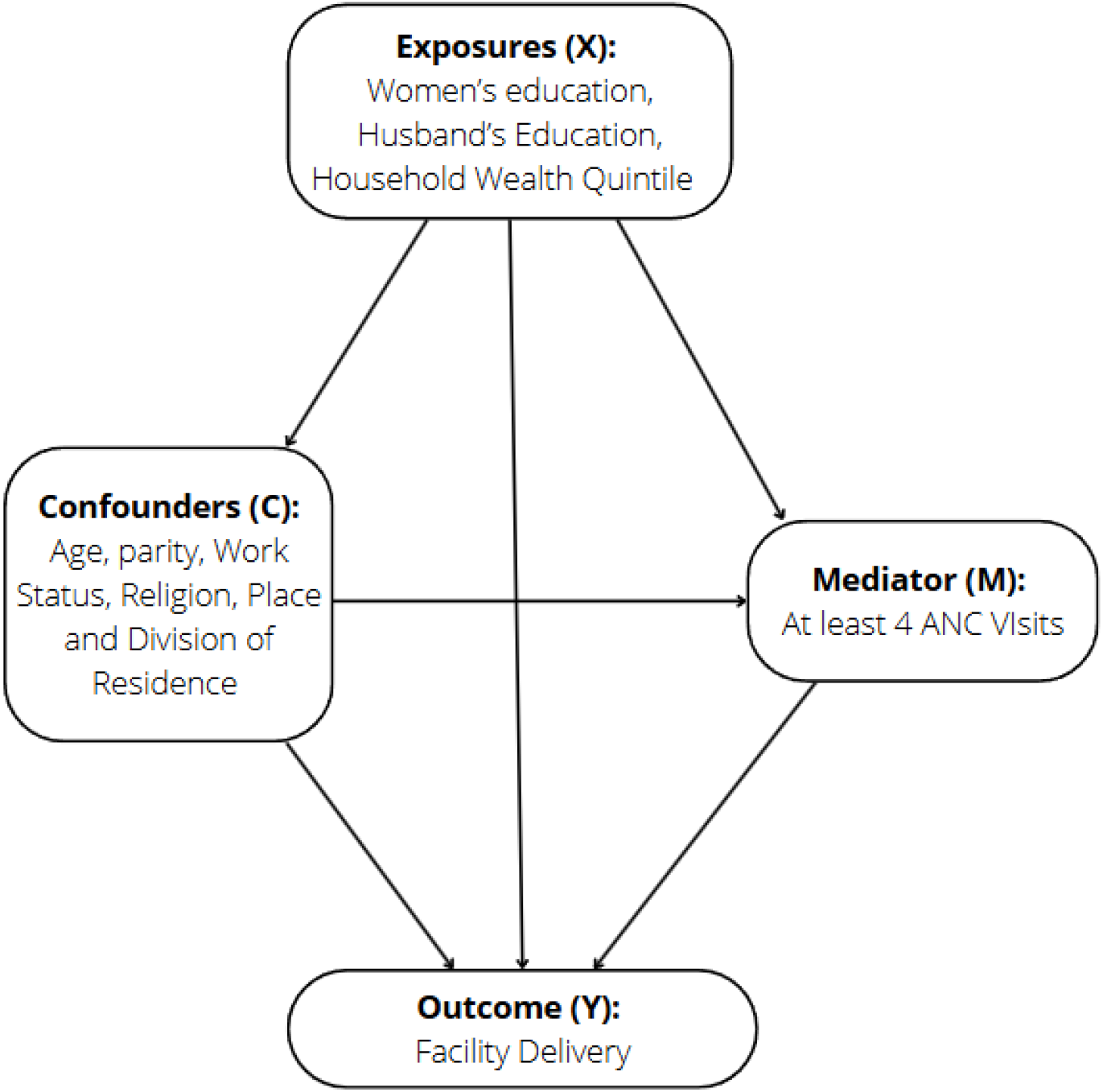
Conceptual Framework Note: Equations to Consider: (1) Exposures (X) and Outcome (Y): Y = X + C;(2) Exposures (X) and Mediator (M): M = X + C; (3) Mediator (M) and Outcome (Y): Y = M + C; and (4) Exposures (X), Mediator (M), and Outcome (Y): Y = M + X + C

#### Exposure Variables

For socioeconomic status, we investigated the following three variables as the exposures (Figure 1): women’s education, husband’s education, and household wealth quintile. Women reported their education, husbands’ education, and whether they were working during the survey period. Women and their husbands’ educational attainment was divided into four categories: no formal education, primary (i.e., one to five years), secondary (i.e., six to ten years), and college or higher (i.e., eleven or more years). The household wealth index score was used to determine the quintiles of household wealth: poorest, poorer, middle, richer, and richest. Principal component analyses of water sources, sanitary facilities, power, basic household construction materials, and household possessions were used to determine this score [16].

#### Other Covariates

Based on published studies, scientific plausibility, and dataset structure, we used the following variables as the potential covariates: maternal age, parity, current work status of the women, place of residence, and division of residence. Three age groups for mothers were identified: 15–24, 25–34, and 35–49 years. Parity was divided into two categories: “primi” (i.e., first pregnancy) and “second or more.” Women who lived in a municipal or city corporation were classified as urban; all other women were classified as rural. Dhaka, Chittagong, Rajshahi, Khulna, Barisal, Rangpur, Sylhet, and Mymensingh were the eight divisions of Bangladesh during the survey period [16].

### Statistical Analysis

First, respondents’ sociodemographic and socioeconomic traits were described according to the presence of at least four ANC visits and facility delivery. The mean and standard errors (SE) were used to report continuous variables. On the other hand, weighted frequencies (n) and percentages (%) were used to report categorical variables. To compare the distribution, Student’s t-tests and Rao Scott’s chi-square tests were used for continuous and categorical variables, respectively.

To test the association of exposure, mediator, and outcome, several equations (exposure-mediator, exposure-outcome, mediator-outcome, and exposure-mediator-outcome) have to be tested together (Figure 1). As the primary mediator and outcomes were binary and the potential exposures were categorical, to test all of the relationships together, generalized structural equation modeling (GSEM) with path analysis was used. Only variables that were significant in bivariate analyses were adjusted into GSEM. All the model fit statistics along with the proportion of mediation, and total, direct, and indirect effects were reported. SEM is a multivariate statistical analysis method that examines the structural and measurement correlations between several variables. Here, all the variables were observed. SEM is an extension of the well-known linear and logistic regression models, incorporating indirect relationships between variables in addition to the direct relationships found in regression models. GSEM is a framework that combines the strengths of SEM and the generalized linear model.

Stata 14.0 was used for the analysis (College Station, TX, USA). The given sample weights and the hierarchical structure of BDHS were considered.

## Results

A total of 4950 women were included in the analysis (Table 1). The mean age of them was 25.7 years (SE: 0.1). The proportion of women with formal education or formally educated husbands was 94.6% and 84.9%, respectively. More than two-thirds of the women were from rural areas, 73.2%, and about 24.4% of them were from Dhaka Division. When we compared women according to the presence of at least four ANC visits, we observed that a majority of the women with at least 4 ANC visits had higher education, higher educated husbands, urban residence, or richer/richest wealth quintiles than women without at least four ANC visits. When we compared women according to hospital delivery, similar to at least 4 ANC visits, women with hospital birth had higher education, higher educated husbands, urban residents, or richer/richest wealth quintiles than women without hospital birth.

**Table 1:**
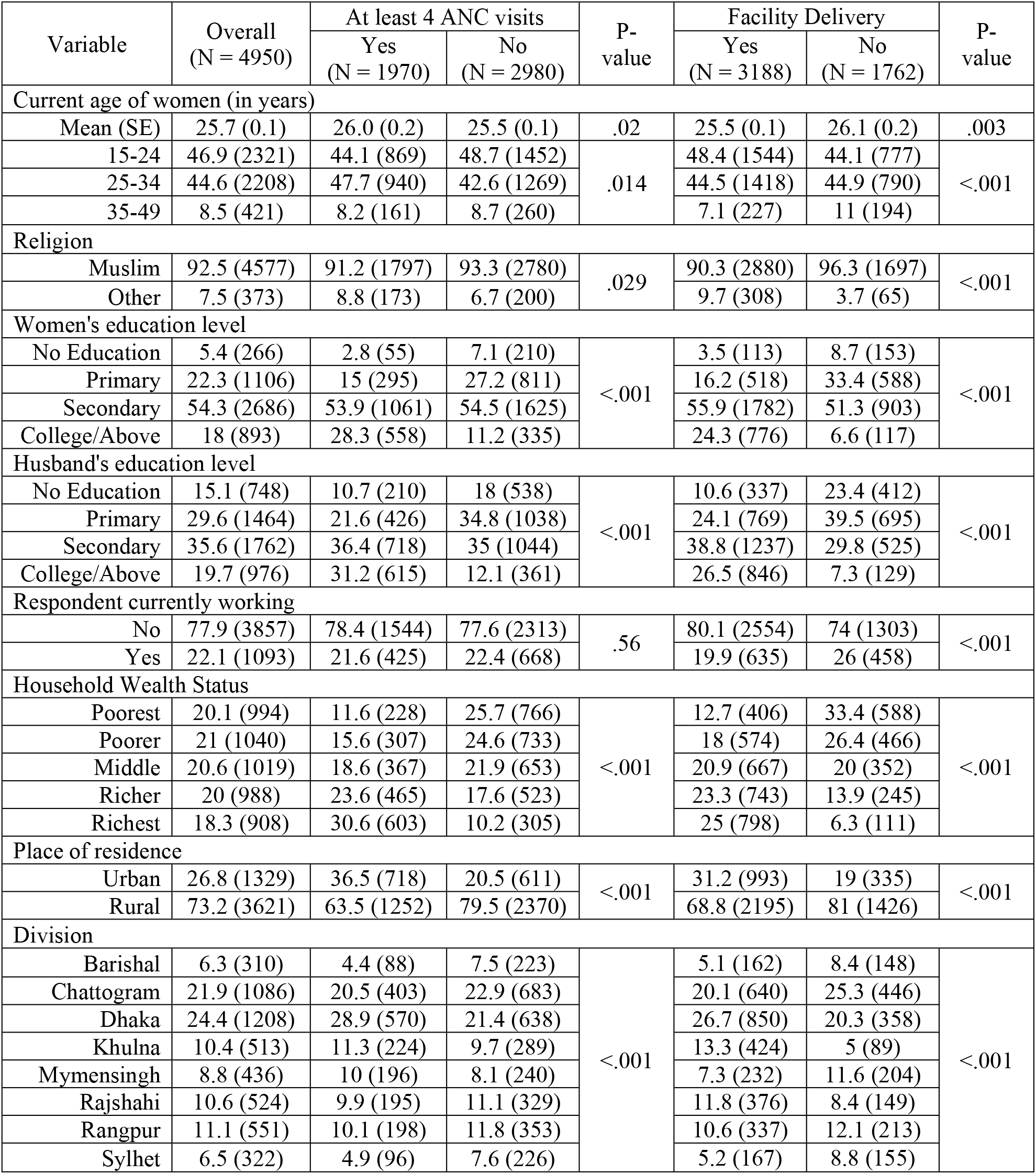
Background characteristics of the study sample according to at least 4 ANC visits and facility delivery.

The proportions of women with at least 4 ANC visits and facility delivery were 39.8% (95% CI: 37.6%-42.0%) and 64.4% (95% CI: 62.1%-66.7%), respectively (Figure 2). The proportion of women who had facility delivery was 81.1% (95% CI: 78.7%-83.3%) and 53.4% (95% CI: 50.6%-56.2%) among women with and without at least 4 ANC visits, respectively. The proportion of at least 4 ANC visits and facility delivery also differed by all three socioeconomic variables.

**Figure 2:**
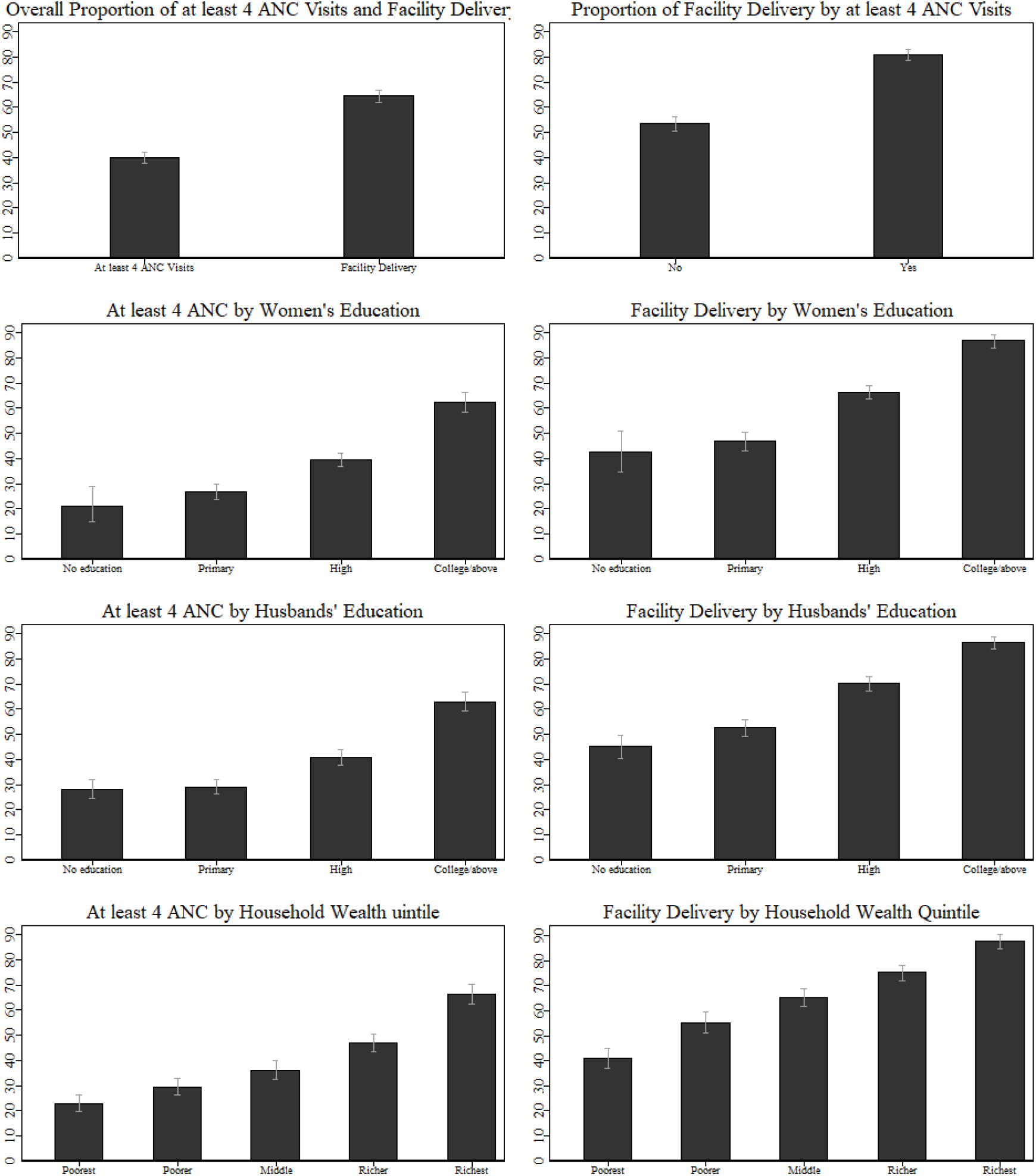
Proportion (%) of Women with at least 4 Antenatal Care Visits and Facility Delivery.

### 1. Adjusted for women’s age, religion, work status, place and division of residence

In Table 2, we reported the association of socioeconomic factors with the variables, ‘at least 4 ANC visits’ and ‘hospital delivery’, adjusted for other variables (e.g., women’s age, religion, work status, place and division of residence). Overall, all three factors (i.e., women’s education, husband’s education, and household wealth quintiles) had significant associations with both variables. For instance, compared to women with the poorest wealth quintile, those with poorer (adjusted odds ratio (AOR): 1.26, 95% confidence interval (CI):1.02-1.55), middle (AOR: 1.43, 95% CI: 1.16-1.77), richer (AOR: 1.98, 95% CI: 1.59-2.45), and richest (AOR: 3.12, 95% CI: 2.45-3.99) wealth quintiles higher odds of at least 4 ANC visits. Next, compared to women with the poorest wealth quintile, those with poorer (AOR: 1.44, 95% CI: 1.19-1.75), middle (AOR: 1.82, 95% CI: 1.48-2.24), richer (AOR: 2.48, 95% CI: 1.98-3.10), and richest (AOR: 3.90, 95% CI: 2.94-5.18) wealth quintiles had higher odds of hospital delivery. Furthermore, women who had at least 4 ANC visits, had more than two and a half times higher odds of hospital delivery (AOR: 2.56, 95% CI: 2.20-2.97).

**Table 2:** Association of socioeconomic variables with at least four ANC visits and facility delivery.

| Variables | At least four ANC visits |  | Facility delivery |  |
| --- | --- | --- | --- | --- |
|  | AOR (95% CI)<sup>1</sup> | P-values | AOR (95% CI)<sup>1</sup> | p-values |
| At least four ANC, Ref: No | --- | -- | 2.56 (2.20,2.97) | <.001 |
| Women's education level, Ref: No formal education |  |  |  |  |
| Primary | 1.36 (0.97,1.92) | .079 | 1.08 (0.79,1.47) | .63 |
| Secondary | 2.04 (1.45,2.87) | <.001 | 1.40 (1.03,1.91) | .032 |
| College/Above | 2.91 (1.98,4.26) | <.001 | 2.38 (1.62,3.51) | <.001 |
| Husbands' education level, Ref: No formal education |  |  |  |  |
| Primary | 0.98 (0.79,1.21) | .82 | 1.11 (0.91,1.35) | .32 |
| Secondary | 1.17 (0.94,1.45) | .17 | 1.46 (1.18,1.81) | <.001 |
| College/Above | 1.75 (1.35,2.28) | <.001 | 2.10 (1.56,2.82) | <.001 |
| Household wealth quintile, Ref: Poorest |  |  |  |  |
| Poorer | 1.26 (1.02,1.55) | .029 | 1.44 (1.19,1.75) | <.001 |
| Middle | 1.43 (1.16,1.77) | .001 | 1.82 (1.48,2.24) | <.001 |
| Richer | 1.98 (1.59,2.45) | <.001 | 2.48 (1.98,3.10) | <.001 |
| Richest | 3.12 (2.45,3.99) | <.001 | 3.90 (2.94,5.18) | <.001 |

Abbreviations: ANC: Antenatal care; AOR: Adjusted odds ratio, CI: Confidence interval

Lastly, we obtained the indirect, direct, and total effects of socioeconomic variables (Table 3). At least 4 ANC had significant mediating effects on the relationship between any of the socioeconomic variables and hospital delivery. For instance, the coefficients of indirect, direct, and total effects of women’s education were 1.96 (95% CI: 0.96-2.96), 1.28 (95% CI: 0.36-2.21), and 3.24 (95% CI: 1.89-4.59), respectively; the proportion of total effects of women’s education on hospital delivery that is mediated through at least 4 ANC visits was 60.5%. On the other hand, the proportions of mediating effects of at least 4 ANC visits on husbands’ education and household wealth quintiles were 34.6% and 41.1%, respectively.

**Table 3:** Indirect and total effects of socioeconomic variables on the relationship of ANC and facility delivery.

| Variable | Indirect Effect |  | Direct Effect |  | Total Effect |  | % of Mediation |
| --- | --- | --- | --- | --- | --- | --- | --- |
|  | Coef. (95% CI) | p-value | Coef. (95% CI) | p-value | Coef. (95% CI) | p-value |  |
| Women's education | 1.96<br>(0.96,2.96) | <0.001 | 1.28<br>(0.36,2.21) | 0.007 | 3.24 (1.89,4.59) | <0.001 | 60.5% |
| Husbands' education | 0.67<br>(0.09,1.26) | 0.029 | 1.22<br>(0.62,1.83) | <0.001 | 1.87 (1.03,2.70) | <0.001 | 34.6% |
| Household Wealth Quintile | 2.26<br>(1.49,3.04) | <0.001 | 3.23<br>(2.53,3.95) | <0.001 | 5.50 (4.46,6.54) | <0.001 | 41.1% |

## Discussion

In this study, we observed that more than half of the women did not receive at least 4 ANC visits and more than one-third of women did not give birth to a health facility. We also found a positive relationship of higher socioeconomic status with these two health utilization variables, and at least four ANC visits partially mediated the relationship between socioeconomic variables and hospital delivery. To our knowledge, this is the first epidemiological study that reported the mediating impact of ANC visits on the relationship of socioeconomic status and at least four ANC visits in Bangladesh.

In Bangladesh and comparable LMICs, socioeconomic differences in maternal and child mortality, as well as in accessing and receiving various maternal and child care services [13,21–23], are reflected in the association between socioeconomic status and this study’s results. Because they are unaware of the advantages of ANC during pregnancy, women with lower educational attainment or spouses with lower educational attainment may not receive proper healthcare services, including at least 4 ANC visits, which ultimately resulted in not having the delivery in the hospital ^20,25,26^. Additionally, because they cannot afford the accompanying fees, women from lower-income families may encounter obstacles while trying to get ANC visits [19,20]. The likelihood of positive outcomes is positively correlated with wealth quintiles and educational attainment. Because people with higher levels of education typically earn more money than those with lower levels of education; furthermore, education, and wealth/income are related [20,21]. Despite the low cost of healthcare services in Bangladesh’s public hospitals, these inequities may be exacerbated by the expenses of travel and hospital stay costs [16].

These findings have ramifications for Bangladesh and other comparable LMICs. Although ANC is the first step in the continuum of care during pregnancy [24], most women did not receive the recommended number or quality of ANC, setting them up for adverse obstetric outcomes in the earliest stage of pregnancy. Future initiatives should address other obstacles to obtaining health treatments, as ANC plays a critical role in lowering mother and infant mortality [10,14]. This study is distinctive in that it looks at the quantity of ANC, showing how it can potentially impact future facility delivery utilization. It is also necessary to consider the identified socioeconomic differences. To improve the situation of maternal and child health, authorities in Bangladesh could therefore address all of the elements that have been identified. Furthermore, a previous study reported that only 18% of women received high-quality ANC, despite 47% receiving it. To put it another way, just one out of three mothers who had four ANC visits obtained high-quality ANC, and just about 37% of participants knew about warning indications [22]. The full benefits of ANC cannot be realized unless its quality is guaranteed. Although we did not look into the quality of health centers where women go for treatment, it can also affect the quality of ANC or facility delivery, as well as the willingness of women to go for future treatment [15,16,21].

This study has some noteworthy strengths. First, the results were generalizable due to the large sample size and the fact that it covered both urban and rural areas in every administrative division. Furthermore, the survey had a high response rate and few missing data points, which guaranteed the accuracy of the findings. Additionally, the study used criteria, questionnaires, and methodologies that were established and validated, allowing comparisons between women from different sociodemographic and socioeconomic backgrounds [16].

It is also necessary to take into account the limitations of the current study. First, we were unable to investigate the relationship between certain elements, including health center quality. Furthermore, memory bias may be introduced by including pregnancy history from the previous three years in this study. It is crucial to remember that this study is cross-sectional, which makes it more difficult to prove causation because there is no temporal certainty [16].

## Conclusion

We examined socioeconomic disparities in ANC utilization and how this can impact hospital delivery utilization in Bangladesh. Considering the lower utilization of at least 4 ANC visits and its mediating impact on the relationship between socioeconomic status and facility delivery, more community-based programs are required to increase awareness and reduce disparities about it. While reducing these disparities may take a long time, would be crucial to improve maternal and child health in the country.

## Data Availability

Data is available from: https://dhsprogram.com/data/available-datasets.cfm

## Acknowledgment

The authors thank the survey participants for their time.

## Consent for publication

Not applicable

## Availability of data and material

Data is available from https://dhsprogram.com/data/available-datasets.cfm

## Ethics Declaration

The ethical approval for the BDHS was provided by institutional review boards (IRB) of ICF International and Bangladesh Medical Research Council. The dataset is available for scientific and academic use upon approval from ICF international.

## Ethics, Consent to Participate, and Consent to Publish declarations

Not applicable.

## Authors contributions

**Conceptualization:** Gulam Muhammed Al Kibria, Zabir Hasan

**Data Cleaning and Analysis:** Gulam Muhammed Al Kibria

**Investigation:** Gulam Muhammed Al Kibria, Zabir Hasan, Md. Shajedur Rahman Shawon, Mohammad Nurunnabi

**Methodology:** Gulam Muhammed Al Kibria

**Writing-Original Draft:** Gulam Muhammed Al Kibria

**Writing – Review & Editing:** Zabir Hasan, Md. Shajedur Rahman Shawon, Mohammad Nurunnabi

## References

1. World Health Organization, United Nations Children Fund, United Nations Population Fund, World Bank, United Nations Population Division. Maternal mortality: Levels and trends 2000 to 2020. 2023. Available: https://iris.who.int/bitstream/handle/10665/366225/9789240068759-eng.pdf?sequence=1

2. UN Inter-Agency Group for Child Mortality Estimation. Levels & Trends in Child Mortality Report 2023. 2023. Available: https://www.unicef.org/publications/files/Child_Mortality_Report_2017.pdf

3. United Nations. Millennium Development Goals. 2000. Available: https://sdgs.un.org/#goal_section

4. Kuhnt J, Vollmer S. Antenatal care services and its implications for vital and health outcomes of children: evidence from 193 surveys in 69 low-income and middle-income countries. BMJ Open. 2017;7:e017122. doi:10.1136/bmjopen-2017-017122

5. Mcdonagh M. Is antenatal care effective in reducing maternal morbidity and mortality? Health Policy Plan. 1996;11: 1–15. doi:10.1093/heapol/11.1.1

6. World Health Organization. WHO recommendations on antenatal care for a positive pregnancy experience. 2016. Available: https://apps.who.int/iris/bitstream/handle/10665/250796/97892415?sequence=1

7. World Health Organization, United Nations Children Fund, United Nations Population Fund, World Bank, United Nations Population Division. Maternal mortality: Levels and trends 2000 to 2017. 2019. Available: https://www.who.int/reproductivehealth/publications/maternal-mortality-2000-2017/en/

8. Lawn JE, Cousens S, Zupan J. 4 million neonatal deaths: When? Where? Why? The Lancet. 2005;365: 891–900. doi:10.1016/S0140-6736(05)71048-5

9. Arroyave L, Saad GE, Victora CG, Barros AJD. Inequalities in antenatal care coverage and quality: an analysis from 63 low and middle-income countries using the ANCq content-qualified coverage indicator. Int J Equity Health. 2021;20: 102. doi:10.1186/s12939-021-01440-3

10. Carroli G, Villar J, Piaggio G, Khan-Neelofur D, Gülmezoglu M, Mugford M, et al. WHO systematic review of randomised controlled trials of routine antenatal care. The Lancet. 2001;357: 1565–1570. doi:10.1016/S0140-6736(00)04723-1

11. Carvajal–Aguirre L, Mehra V, Amouzou A, Khan SM, Vaz L, Guenther T, et al. Does health facility service environment matter for the receipt of essential newborn care? Linking health facility and household survey data in Malawi. J Glob Health. 2017;7: 020508. doi:10.7189/jogh.07.020508

12. Carlough M, McCall M. Skilled birth attendance: What does it mean and how can it be measured? A clinical skills assessment of maternal and child health workers in Nepal. Int J Gynecol Obstet. 2005;89: 200–208. doi:10.1016/j.ijgo.2004.12.044

13. Khanam R, Baqui AH, Syed MIM, Harrison M, Begum N, Quaiyum A, et al. Can facility delivery reduce the risk of intrapartum complications-related perinatal mortality? Findings from a cohort study. J Glob Health. 2018;8: 010408. doi:10.7189/jogh.08.010408

14. Tura G, Fantahun M, Worku A. The effect of health facility delivery on neonatal mortality: systematic review and meta-analysis. BMC Pregnancy Childbirth. 2013;13: 18. doi:10.1186/1471-2393-13-18

15. National Institute of Population Research and Training (NIPORT), Mitra and Associates, ICF International. Bangladesh Demographic and Health Survey 2017-18. Dhaka, Bangladesh; 2020.

16. National Institute of Population Research and Training (NIPORT), Mitra and Associates, ICF International. Bangladesh Demographic and Health Survey 2022. Dhaka, Bangladesh; 2024.

17. National Institute of Population Research and Training (NIPORT), Mitra and Associates, ICF International. Bangladesh Demographic and Health Survey 1993-94. Dhaka, Bangladesh; 2013. Available: https://dhsprogram.com/pubs/pdf/fr265/fr265.pdf

18. Kibria GMA, Burrowes V, Choudhury A, Sharmeen A, Ghosh S, Kalbarczyk A. A comparison of practices, distributions and determinants of birth attendance in two divisions with highest and lowest skilled delivery attendance in Bangladesh. BMC Pregnancy Childbirth. 2018;18. doi:10.1186/s12884-018-1770-9

19. Bohren MA, Hunter EC, Munthe-Kaas HM, Souza JP, Vogel JP, Gülmezoglu AM. Facilitators and barriers to facility-based delivery in low- and middle-income countries: a qualitative evidence synthesis. Reprod Health. 2014;11: 71. doi:10.1186/1742-4755-11-71

20. Nayeem J, Stennett C, Sharmeen A, Hossain MM, Kibria GMA. Rural-urban differences in distributions and determinants of facility delivery among women in Bangladesh. Glob Health J. 2023;7: 222–229. doi:10.1016/j.glohj.2023.12.001

21. Kibria GMA, Crispen R. Disparities, distribution, and determinants in appropriate timely initiation, number, and quality of antenatal care in Bangladesh: Evidence from Demographic and Health Survey 2017–18. Abdo RA, editor. PLOS Glob Public Health. 2023;3: e0002325. doi:10.1371/journal.pgph.0002325

22. Kibria GMA, Ghosh S, Hossen S, Barsha RAA, Sharmeen A, Uddin SMI. Factors affecting deliveries attended by skilled birth attendants in Bangladesh. Matern Health Neonatol Perinatol. 2017;3. doi:10.1186/s40748-017-0046-0

23. Shahjahan M, Chowdhury HA, Al-Hadhrami AY, Harun GD. Antenatal and postnatal care practices among mothers in rural Bangladesh: A community based cross-sectional study. Midwifery. 2017;52: 42–48. doi:10.1016/j.midw.2017.05.011

24. Singh K, Story WT, Moran AC. Assessing the Continuum of Care Pathway for Maternal Health in South Asia and Sub-Saharan Africa. Matern Child Health J. 2016;20: 281–289. doi:10.1007/s10995-015-1827-6

